# Ethnic variation in outcome of people hospitalised with Covid-19 in Wales (UK): A rapid analysis of surveillance data using Onomap, a name-based ethnicity classification tool

**DOI:** 10.1101/2020.06.22.20136036

**Authors:** Daniel Rh Thomas, Oghogho Orife, Amy Plimmer, Christopher Williams, George Karani, Meirion R Evans, Paul A Longley, Janusz Janiec, Roiyah Saltus, A Giri Shankar

## Abstract

There is growing evidence that ethnic minorities in Europe are disproportionately affected by Covid-19. Using a name-based ethnicity classifier, we found that hospitalised Black, Asian and minority ethnic cases were younger and more likely to be admitted to intensive care (ICU). Pakistani, Bangladeshi and White - other than British or Irish, ethnic groups were most at risk. In this study, older age and male gender, but not ethnicity, were associated with death in hospitalised patients.

## Manuscript

There is growing evidence that Black, Asian and other minority ethnic (BAME) people living in Europe are at increased risk of infection with SARS-CoV-2 and, if infected, are more likely to have severe disease.^1^ In the United Kingdom, the Intensive Care National Audit and Research Centre first raised concerns that BAME people were over-represented amongst Covid-19 patients admitted to intensive care.^2^ These findings were reported widely in the media and discussed in opinion pieces.^3-7^ In Wales, the First Minister established an advisory group to examine the issue and provide recommendations to reduce ethnic inequality in Covid-19 outcomes.^8^

Investigating ethnic health inequalities is hampered by poor recording of ethnicity in clinical data. This is the case for Covid-19 notifications and laboratory reports in Wales. In order to rapidly investigate ethnic variation in Covid-19 epidemiology, we applied Onomap, a name-based ethnicity classification tool developed by the Department of Geography at University College London,^9^ to routinely collected, named Covid-19 laboratory test data, held by Public Health Wales Communicable Disease Surveillance Centre.

We used individual person data on: (1) 35,618 SARS-CoV-2 PCR tests carried out by Public Health Wales and authorised as at 1300 hrs, 3 May 2020 from Microbiology Datastore, a repository of test results recorded in the all-Wales Laboratory Information Management System; (2) 3,394 hospitalised patients (people admitted to hospital within 14 days of a positive SARS-CoV-2 test or individuals who tested positive for SARS-CoV-2 whilst in hospital) as at 1700 hrs, 3 May 2020 extracted from IC-Net, an infection prevention and control information management system; and (3) 1,071 Covid-19 in-hospital deaths (Covid-19 cases who died in hospital, and had a positive test result for SARS-CoV-2 28 days or less than the date of death or 7 days after death) in all reported cases hospitalised to 1700hrs, 3 May, as at 31 May 2020. Ethnicity was categorised using Onomap and the 2001 Census classification of ethnicity.^10^ We collapsed these categories further into: ‘White British or Irish’, ‘White Other’, ‘Asian or British Asian’, ‘Black or Black British’, ‘other ethnicity’ and ‘unclassified’, with a further aggregation to create a ‘BAME’ field, containing all ethnicities other than ‘White British’, ‘White Irish’, or ‘White Other’. Unclassified observations were excluded.

Using the cohort of 3,394 hospitalised patients, we carried out a logistic regression to calculate odds ratios for the outcomes: (a) admitted to intensive care and (b) mortality, with 95% confidence intervals, for ethnic groups, in each case using White British or Irish ethnicity as the baseline comparator. Independent variables were gender and age group. Multivariable analyses were then used to calculate odds ratios for ethnic groups whilst controlling for sex and age group. Differences in the distributions of previously reported risk factors for fatal outcomes (age, gender, medical history)^11^ were investigated further in White and BAME groups. The Mann Whitney two-sample test was used to compare differences in the age distribution of BAME and White deaths. Odds ratios with 95% confidence intervals were calculated to compare proportion male and proportion with underlying health conditions amongst deceased BAME and White individuals. All analysis was carried out using Stata 14.^12^

## Findings

Onomap estimated the ethnicity of 98.9% (10,413/10,524) of tested individuals, 99.6% (3,382/3,394) of those hospitalised, 99.3% (272/274) of those admitted to intensive care, and 99.8% (1,069/1,071) of those who died following admission to hospital.

By classifying ethnicity using names, we estimate that 4.9% (n=1759) tests were of Black, Asian and other minority (BAME) individuals (Table 1). Using the most recent Statistics Wales population estimates for ethnic groups in Wales,^13^ this represents 960 tests per 100, 000 population in BAME, compared to 1,145 tests per 100,000 population in White ethnic groups.

**Table 1.**
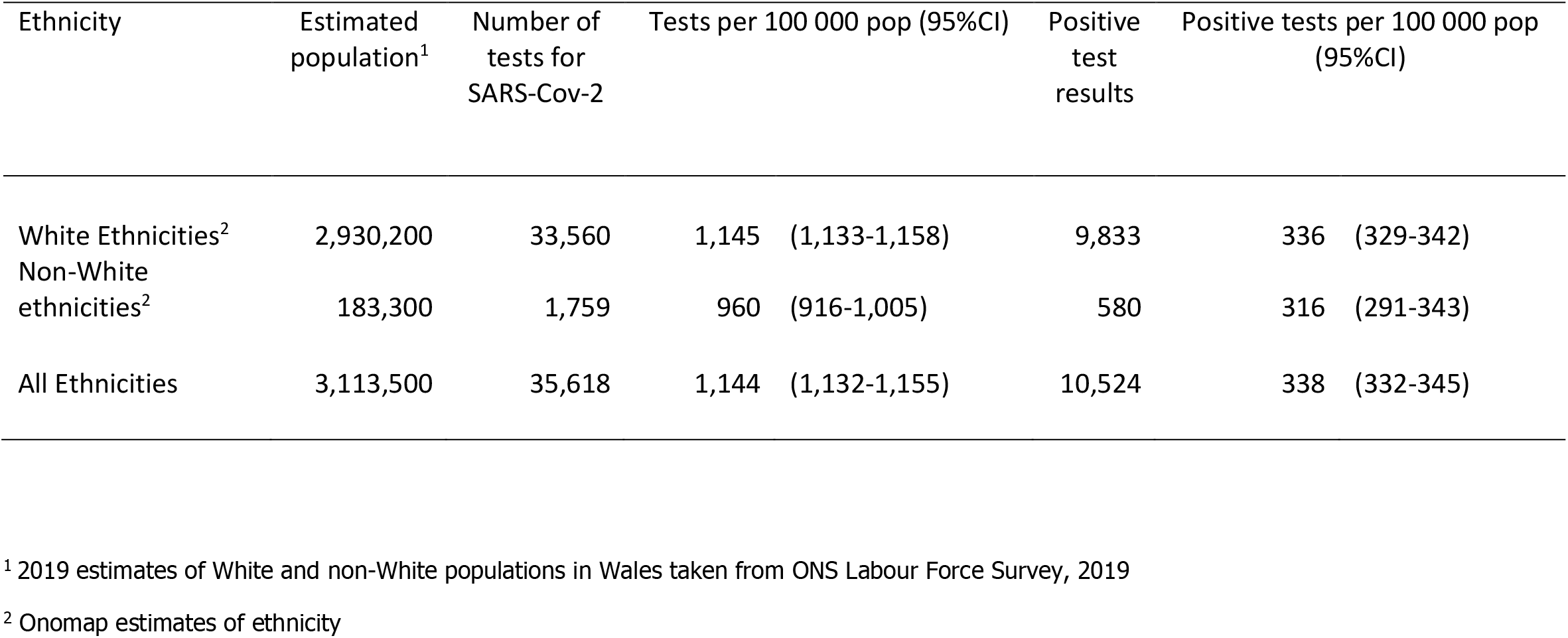
**Proportions of the population tested for SARS-CoV-2 in Wales and associated proportions of positive test results, White and BAME individuals**

| Ethnicity | Estimated population<sup>1</sup> | Number of tests for SARS-Cov-2 | Tests per 100 000 pop (95%CI) | Positive test results | Positive tests per 100 000 pop (95%CI) |
| --- | --- | --- | --- | --- | --- |
| White Ethnicities<sup>2</sup> | 2,930,200 | 33,560 | 1,145 (1,133-1,158) | 9,833 | 336 (329-342) |
| Non-White ethnicities<sup>2</sup> | 183,300 | 1,759 | 960 (916-1,005) | 580 | 316 (291-343) |
| All Ethnicities | 3,113,500 | 35,618 | 1,144 (1,132-1,155) | 10,524 | 338 (332-345) |

<sup>1</sup> 2019 estimates of White and non-White populations in Wales taken from ONS Labour Force Survey, 2019
<sup>2</sup> Onomap estimates of ethnicity

Of 10,524 people tested positive for SARS-CoV-2 in Wales to 3 May 2020, Onomap classified 9,833 in White ethnic groups and 580 in BAME groups. Proportions with positive test results were similar for both groups: 336 per 100,000 of the White group tested positive and 316 per 100,000 in the BAME group. Trends in those tested positive should be interpreted with caution as they most likely reflect testing policy as well as incidence. Of all those testing positive, a smaller proportion (18.1%) of those tested in the BAME group attended hospital compared to the White group (33.4%: see Table 2). However, the trend was reversed in people aged 50 to 59 years: 26.4% of positive BAME individuals aged 50-59 years attended hospital, compared to 19.0% of White individuals testing positive. The median age of hospitalised BAME individuals was 51 years compared to 75 years for White individuals (p<0.01; Mann Whitney 2 sample test).

**Table 2.** **Ethnicity breakdown of individuals tested for SARS-CoV-2 in Wales and proportions hospitalised, attending intensive care units (ICU), and deceased**

| Ethnicity<sup>1</sup> | Positive tests | Hospitalised | % hospitalised (95%CI) |  | Attending ICU | % hospitalised attending ICU (95% CI) |  | Deceased citing Covid-19 | % hospitalised and deceased citing Covid-19 (95%CI) |  |
| --- | --- | --- | --- | --- | --- | --- | --- | --- | --- | --- |
| All ethnicities | 10,524 | 3,394 | 32.3 | (31.4-33.1) | 274 | 8.0 | (7.2-9.0) | 1,071 | 31.6 | (30.0-33.1) |
| White British and Irish | 9,439 | 3,194 | 34.0 | (32.8-34.8) | 230 | 7.2 | (6.3-8.1) | 1,041 | 32.5 | (31.0-34.2) |
| White – other | 394 | 83 | 21.1 | (17.0-25.1) | 21 | 25.3 | (16.0-34.7) | 18 | 21.7 | (12.8-30.5) |
| **All white ethnicities** | **9,833** | **3,277** | **33.4** | **(32.4-34.3)** | **251** | **7.6** | **(6.7-8.6)** | **1,059** | **32.3** | **(30.7-33.9)** |
| Asian and British Asian (Indian, Pakistani, Bangladeshi) | 289 | 61 | 21.1 | (16.4-25.8) | 17 | 27.9 | (16.6-39.1) | <10 | - |  |
| Black and Black British | 42 | <10 | - |  | <10 | - |  | <10 | - |  |
| Chinese | 32 | <10 | - |  | <10 | - |  | <10 | - |  |
| Other | 217 | 30 | 13.8 | (9.2-18.4) | <10 | - |  | <10 | - |  |
| **All non-white ethnicities** | **580** | **105** | **18.1** | **(15.0-21.2)** | **21** | **20.0** | **(12.3-27.7)** | **10** | **9.5** | **(3.9-15.1)** |
| Unknown | 111 | 12 | 10.8 | (5.0-16.6) | 2 | 16.7 | (0.4-37.7) | 2 | 16.6 | (0.4-37.7) |

<sup>1</sup> Onomap estimates of ethnicity

Of those attending hospital, a much higher proportion (20.0%) of BAME individuals were admitted to intensive care compared to White individuals (7.7%). Proportions of hospitalised patients admitted to intensive care (ICU) were highest amongst the ‘Asian and British Asian-Indian, Pakistani and Bangladeshi’ (27.9%) and ‘White – other’ (25.3%) groups. The median age of BAME patients admitted to ICU was 51 years compared to 58 years for White individuals (p=0.02; Mann Whitney 2 sample test). Amongst hospitalised patients aged between 50-59 years, 25% of BAME patients were admitted to ICU compared to 19.5% of White patients. More patients died in hospital without being admitted to ICU. Of all those attending hospital, 9.5% of patients identified as BAME died compared to 32.3% of White patients (Table 2).

We successfully linked all records of 3,394 people hospitalised with Covid-19, those admitted to ICU, and those who died in hospital, all as at 3 May 2020, using NHS numbers. Intensive care was more likely in hospitalised males (aOR: 1.92, 95% CI: 1.46-2.53) and in younger patients (Table 3, Figure 1). When specific ethnicities were examined, being admitted to ICU was more likely in ‘White Other’, ‘Asian and British Asian – Bangladeshi’, ‘Asian and British Asian – Indian’, and ‘Asian and British Asian – Pakistani’ ethnic groups. After adjusting for gender and age, ‘White Other’ (aOR: 2.54, 95% CI: 1.47-4.37), ‘Bangladeshi’(aOR: 6.57, 95%CI: 1.02-42.17), ‘Pakistani’ (aOR: 2.25, 95%CI: 1.05-4.79) ethnic groups remained significantly more likely to be admitted to ICU.

**Table 3.** **Personal characteristics associated with severe outcomes for Welsh residents hospitalised with Covid-19, to 3 May 2020**

| Personal characteristic |  | Hospitalised | Received intensive care |  |  |  | Died |  |  |
| --- | --- | --- | --- | --- | --- | --- | --- | --- | --- |
|  |  |  | Univariate analysis |  | Multivariate analysis |  | Univariate analysis |  | Multivariate analysis |
|  |  |  | OR (95% CI) |  | OR (95% CI) |  | OR (95% CI) |  | OR (95% CI) |
| Gender |  |  |  |  |  |  |  |  |  |
|  | Female | 1576 | - |  | - |  | - |  | - |
|  | Male | 1818 | **1.86 (1.43-2.42)** |  | **1.92 (1.46-2.53)** |  | **1.33 (1.15-1.54)** |  | **1.35 (1.15-1.57)** |
| Age |  |  |  |  |  |  |  |  |  |
|  | 0-49 | 420 | - |  | - |  | - |  | - |
|  | 50-59 | 429 | 1.21 (0.86-1.71) |  | 1.22 (0.85-1.76) |  | **2.38 (1.43-3.97)** |  | **2.23 (1.34-3.73)** |
|  | 60-69 | 482 | 0.71 (0.49-1.03) |  | 0.71 (0.48-1.04) |  | **6.20 (3.91-9.95)** |  | **5.73 (3.58-9.17)** |
|  | 70 and over | 2063 | **0.13 (0.09-0.19)** |  | **0.14 (0.09-0.20)** |  | **12.54 (8.16-19.26)** |  | **11.77 (7.62-18.18)** |
| Ethnicity |  |  |  |  |  |  |  |  |  |
|  | White: British | 3,130 | - |  | - |  | - |  | - |
|  | White: Irish | 64 | 0.63 (0.20-2.02) |  | 0.64 (0.19-2.11) |  | 0.81 (0.46-1.40) |  | 0.7 (0.39-1.23) |
|  | White: other | 83 | **4.33 (2.60-7.23)** |  | **2.54 (1.47-4.37)** |  | 0.57 (0.34- 0.97) |  | 1.06 (0.60-1.89) |
|  | Asian and British Asian: Bangladeshi | <10 | **6.39 (1.16-35.10)** |  | **6.57 (1.02-42.17)** |  | 0.41 (0.05-3.53) |  | 0.56 (0.06-5.63) |
|  | Asian and British Asian: Chinese | <10 | 1.83 (0.22-14.91) |  | 1.06 (0.12-9.08) |  | - |  | - |
|  | Asian and British Asian: Indian | 16 | **5.81 (2.00-16.87)** |  | 2.60 (0.87-7.82) |  | 0.48 (0.14-1.67) |  | 1.15 (0.30-4.44) |
|  | Asian and British Asian: Pakistani | 39 | **4.41 (2.12-9.16)** |  | **2.25 (1.05-4.79)** |  | **0.24 (0.08-0.66)** |  | 0.61 (0.20-1.83) |
|  | Black and Black British: African | <10 | 2.56 (0.30-21.99) |  | 1.33 (0.14-12.43) |  | 0.41 (0.05-3.53) |  | 0.61 (0.06-6.01) |
|  | Any other ethnic group | 30 | 0.91 (0.22-3.86) |  | 0.44 (0.10-1.91) |  | **0.07 (0.01-0.52)** |  | 0.15 (0.02-1.11) |
|  | Unknown ethnicity | 12 | 2.56 (0.56-11.74) |  | 1.97 (0.40-9.70) |  | 0.41 (0.09-1.88) |  | 0.71 (0.14-3.57) |

**Figure 1.**
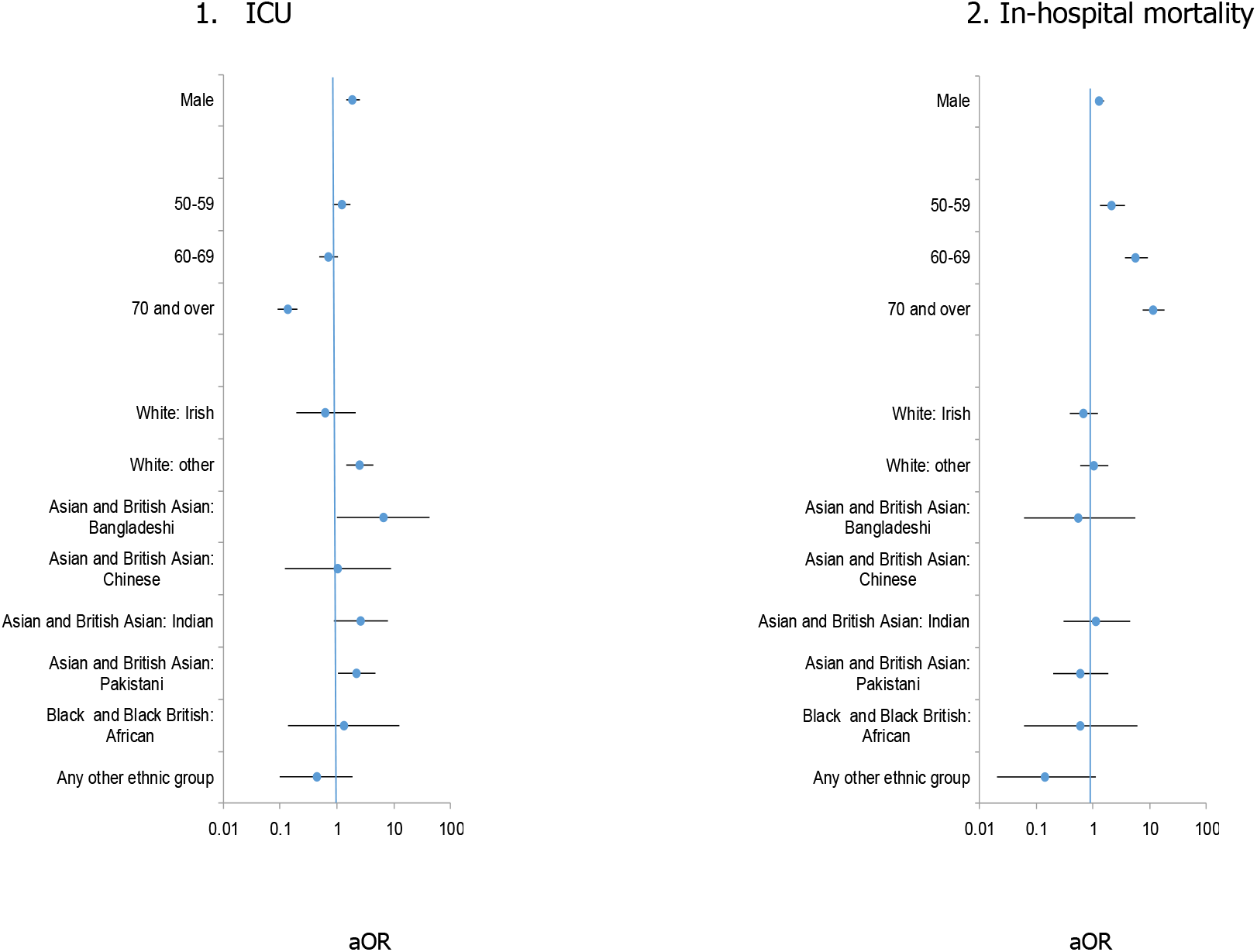
Determinants of: 1. Being admitted to intensive care unit (ICU); and 2. In-hospital mortality in 3,394 individuals hospitalised with Covid-19 in Wales to 3 May 2020, as at 31 May 2020. Adjusted odds ratios (aOR) with 95% confidence intervals are given for male gender, compared to female, older age groups compared to those aged less than 50 years, and Onomap estimated ethnicities, compared to ‘White British’. Odds ratios greater than one represent an increased risk; odds ratios less than one represent a decreased risk. 95% confidence intervals not crossing one reflect that the odds ratio is statistically significant.

Likelihood of dying was significantly higher for hospitalised males. This effect remained after adjusting for age and ethnicity (aOR: 1.35, 95%CI: 1.15-1.57) (Table 3, Figure 1). There was a strong association between increasing age and death from Covid-19 which remained after adjusting for gender and ethnicity (aOR for aged 70 years and over: 11.77 (95% CI: 7.62-18.18). However, there was no evidence from this study that BAME groups were more likely to die from Covid-19 than White-British or Irish groups, even after adjusting for gender and age (Table 3).

To investigate further, we compared the differences in the distribution of previously reported risk factors for fatal outcome^11^ in White and BAME groups who had died. BAME people who died in Wales with Covid-19 were younger than White people who died (BAME median age 74 compared to 80 for White people; p=0.06, Mann-Whitney 2 sample test). Underlying chronic disease was recorded for 48% of deaths. For those that had a medical history recorded, nearly all had an underlying chronic condition that would put them at increased risk of serious Covid-19 symptoms, and there was no difference between White and BAME groups.

## Discussion

This was a rapid initial analysis of existing surveillance data using name-based ethnicity classification software. It adds to the increasing evidence of variation in Covid-19 outcomes in ethnic minorities in Europe. The finding that certain minority ethnic groups are at higher risk of being admitted to intensive care but are no more likely to die than the White British and Irish group was also found in the recent CO-CIN cohort study involving 23,577 Covid-19 patients attending hospitals in the UK.^14^

Onomap has been used widely as a tool in public health, for example in studies investigating variation in influenza mortality,^15^ hepatitis B infection ^16^ and HPV vaccination uptake.^17^ However, Onomap has limitations, and all findings should be interpreted in light of these. We previously validated the tool using data containing self-reported or healthcare professional-reported ethnicity (Supplementary Table 1). Onomap performs well for most ethnicities, but has a low sensitivity for Black or Black British individuals. Risks identified for Black and Black British groups are therefore likely to be underestimated. Kandt and Longley have published a comparison of Onomap with 2011 Census data.^18^

There is an urgent need for all European countries carrying out Covid-19 surveillance to report trends by ethnicity, in order to inform local infection prevention and control policy and practice. Ethnic variation should also be considered in the design of interventions, and in crisis communication. In Wales, an occupational risk assessment tool has been developed with the aim of reducing risk of infection in those most vulnerable to severe infection.^19^ This tool, developed initially for the health care sector, is for all ethnicities, but includes a weighting to account for the emerging evidence of increased risk in BAME individuals.

## Data Availability

Data referred to in the manuscript are not publicly available.

## Acknowlegements

Onomap was purchased from Publicprofiler Ltd. The authors acknowledge the many laboratory and surveillance staff in Public Health Wales involved in developing and maintaining routine Covid-19 surveillance in Wales. Victoria McClure assisted with extracting data from IC-Net.

## Authors’ contributions

Daniel Thomas designed the study, contributed to the analysis and wrote the manuscript. Ogogho Orife contributed to the analysis and commented on the manuscript. Amy Plimmer, George Karani, George Karani, Meirion Evans, Janusz Janiec and Roiyah Saltus commented on the manuscript and contributed to the validation work. Paul Longley commented on the methodology and results, including use of the names classification tool. Chris Williams and Giri Shankar commented on the design and analysis and manuscript.

## Supplementary Material

**Supplementary Table 5.** Validation of Onomap. Estimated sensitivity and specificity of Onomap by ethnic group. Calculated by measuring the performance of Onomap to predict ethnicity in three clinical data sets^1^ already containing self-reported or healthcare professional-reported ethnicity

| Ethnicity | Ethnicity reported by participant | Ethnicity predicted by Onomap | Ethnicity correctly predicted | Sensitivity | Specificity |
| --- | --- | --- | --- | --- | --- |
| White British or Irish | 1681 | 1811 |  |  |  |
| Other White | 3235 | 3418 |  |  |  |
| **Total White** | 4916 | 5229 | 4844 | **98.5%** | **77.7%** |
| Indian | 364 | 239 |  |  |  |
| Pakistani | 313 | 348 |  |  |  |
| Bangladeshi | 96 | 88 |  |  |  |
| Chinese | 55 | 18 |  |  |  |
| Other Asian | 9 | 118 |  |  |  |
| **Total Asian or Asian British** | 837 | 811 | 609 | **72.8%** | **96.5%** |
| Black - African | 344 | 142 |  |  |  |
| Black - Caribbean | 10 | 1 |  |  |  |
| Other Black | 23 | 0 |  |  |  |
| **Total Black or Black British** | 377 | 143 | 112 | **29.7%** | **99.5%** |
| Arabic | 36 | 279 |  |  |  |
| Other | 9 | 4 |  |  |  |
| **Other Ethnic Group** | 45 | 283 | 24 | **53.3%** | **96.1%** |
| **Mixed** | 234 | 0 | - |  |  |
| **Unclassified/Unknown** | 231 | 174 | 8 | **3.5%** | **97.4%** |
| **Total** | 6640 | 6640 | **5589** | **87.4%** | **96.1%** |

<sup>1</sup> Three data sets which included self-reported or healthcare professional-reported ethnicity were used to validate Onomap: A list of individuals attending a mosque in Wales who were offered screening for hepatitis C (n=189), a list of tuberculosis patients notified by doctors in Wales (n=3267) and a list of patients attending an infectious disease clinic in Poland (n=3184).

### Ethical and privacy considerations

Ethical oversight of the project was provided by Public Health Wales NHS Trust R&D Division. As this work was carried out as part of the health protection response to a public health emergency in Wales, using routinely collected surveillance data, Public Health Wales R&D Division advised that NHS research ethics approval was not required. The use of named patient data in the investigation of communicable disease outbreaks and surveillance of notifiable disease is permitted under Public Health Wales’ Establishment Order. Data were held and processed under Public Health Wales’ information governance arrangements, in compliance with the Data Protection Act, Caldicott Principles and Public Health Wales guidance on the release of small numbers. No data identifying protected characteristics of an individual were released outside Public Health Wales. Validation work included in a supplementary table was from a project that had previously received permission from the Confidentiality Advisory Group to process patient data on tests for viral hepatitis carried out by laboratories in Wales, and research ethics approval from West of Scotland REC4 (Application title: Incidence of infectious disease in BME groups using Onomap; CAG reference: 16/CAG/0133; IRAS project ID: 210327 REC reference: 16/WS/018).

## References

1. Pan D, Sze S, Minhas JS, Bangash MN, Pareek N, Divall P et al., The impact of ethnicity on clinical outcomes in COVID-19: A systematic review, EClinicalMedicine. 2020. https://doi.org/10.1016/j.eclinm.2020.100404

2. Intensive Care National Audit and Research Centre. ICNARC report on COVID-19 in critical care [Internet]. 2020. Available from: https://www.icnarc.org/Our-Audit/Audits/Cmp/Reports [accessed 14 June 2020]

3. The Guardian. Ethnic minorities dying of Covid-19 at higher rate, analysis shows. Available from: https://www.theguardian.com/world/2020/apr/22/racial-inequality-in-britain-found-a-risk-factor-for-covid-19 [accessed 14 June 2020].

4. Cifuentes R. All in it together? The impact of Coronavirus on BAME people in Wales. Bevan Foundation, 10 April 2020. Available from: https://www.bevanfoundation.org/commentary/all-in-it-together-the-impact-of-coronavirus-on-bame-people-in-wales/ [accessed 14 June 2020].

5. Pareek M, Bangash MN, Pareek N, Pan D, Sze S, Minhas JS. et al.. Ethnicity and COVID-19: an urgent public health research priority. Lancet 2020. https://www.thelancet.com/journals/lancet/article/PIIS0140-6736(20)30922-3/fulltext

6. BMA. Press release: Review into COVID-19 impact on BAME communities must be backed by real-time data and include measures to address problem now, says BMA. Available from: https://www.bma.org.uk/news-nd-opinion/review-into-covid-19-impact-on-bame-communities-must-be-backed-by-real-time-data-and-include-measures-to-address-problem-now-says-bma [accessed 14 June 2020].

7. NHS Confederation briefing. The impact of COVID-19 on BME communities and health and care staff. Available from: https://www.nhsconfed.org/resources/2020/04/the-impact-of-covid19-on-bme-communities-and-staff [accessed 14 June 2020].

8. Welsh Government. Press release: Wales BAME Covid-19 health advisory group takes a cross-Government approach https://gov.wales/wales-bame-covid-19-health-advisory-group-takes-cross-government-approach[accessed 14 June 2020].

9. Public Profiler. Onomap, name classification software. https://www.publicprofiler.org [accessed 14 June 2020].

10. Office for National Statistics. Ethnic group statistics: A guide for the collection and classification of ethnicity data. 2003. Available from: https://web.archive.org/web/20090419003227/http://www.statistics.gov.uk/about/ethnic_group_statistics/downloads/ethnic_group_statistics.pdf [accessed 14 June 2020].

11. The OpenSAFELY Collaborative, Williamson E, Walker AJ, Bhaskaran KJ, Bacon S, Bates C et al. OpenSAFELY: factors associated with COVID-19-related hospital death in the linked electronic health records of 17 million adult NHS patients. MedRxiv 2020.05.06.20092999; doi: https://doi.org/10.1101/2020.05.06.20092999

12. Stata 14. Available from: https://www.stata.com/stata14/

13. StatsWales. Ethnicity by area and ethnic group. Available from: https://statswales.gov.wales/Catalogue/Equality-and-Diversity/Ethnicity/ethnicity-by-area-ethnicgroup [accessed 14 June 2020].

14. Harrison, E., Docherty, A., Semple C. CO-CIN. Investigating associations between ethnicity and outcome from COVID-19. Report to UK Scientific Advisory Group on Emergencies (SAGE), 25 April 2020. Available from: file:///Y:/OUTBREAKS%202007-2020/2020/2020%20Wuhan%20pneumonia/Ethnicity%20work/Articles/s0 238-co-cin-report-ethnicity-outcomes-250420-sage29.pdf [accessed 14 June 2020].

15. Zhao, H., Harris, R.J., Ellis, J. and Pebody, R.G., 2015. Ethnicity, deprivation and mortality due to 2009 pandemic influenza A(H1N1) in England during the 2009/2010 pandemic and the first post-pandemic season. Epidemiology and infection, 143(16), pp. 3375–3383

16. Binka M, Butt ZA, Wong S, Chong M, Buxton JA, Chapinal N et al. Differing profiles of people diagnosed with acute and chronic hepatitis B virus infection in British Columbia, Canada. World Journal of Gastroenterology, 2018, 24(11):1216–1227

17. Pollock KG, Tait B, Tait J, Bielecki K, Kirolos A, Willocks L, Gorman DR. Evidence of decreased HPV vaccine acceptance in Polish communities within Scotland. Vaccine, 2018, 37(5):690–692 DOI: 10.1016/j.vaccine.2018.10.097

18. Kandt J, Longley PA (2018) Ethnicity estimation using family naming practices. PLoS ONE 13(8): e0201774. https://doi.org/10.1371/journal.pone.0201774. xAvailable from: https://journals.plos.org/plosone/article?id=10.1371/journal.pone.0201774).

19. Welsh Government. All Wales Covid-19 Workforce Risk assessment Tool. Available from: https://gov.wales/sites/default/files/publications/2020-05/covid-19-workforce-risk-assessment-tool.pdf [accessed14 June 2020].

